# Angiotensin-converting enzyme (*ACE1, ACE2*) gene variants are associated with COVID19 severity depending on the hypertension status

**DOI:** 10.1101/2020.06.11.20128033

**Authors:** Juan Gómez, Guillermo M Albaiceta, Marta García-Clemente, Carlos López-Larrea, Laura Amado-Rodríguez, Tamara Hermida, Ana I. Enriquez, Pablo Herrero, Santiago Melón, Marta E. Alvarez-Argüelles, Susana Rojo-Alba, Alvaro Leal-Negredo, Elías Cuesta-Llavona, Victoria Alvarez, Rebeca Lorca, Eliecer Coto

## Abstract

**Background:** The Angiotensin system is implicated in the pathogenesis of COVID19. First, ACE2 is the cellular receptor for SARS-COv-2, and expression of the *ACE2* gene could regulate the individual’s susceptibility to infection. In addition, the balance between ACE1 and ACE activity has been implicated in the pathogenesis of respiratory diseases and could play a role in the severity of COVID19. Functional *ACE1*/*ACE2* gene polymorphisms have been associated with the risk of cardiovascular and pulmonary diseases, and could thus also contribute to the outcome of COVID19.

**Methods:** We studied 204 COVID19 patients (137 non-severe and 67severe-ICU cases) and 536 age-matched controls. The *ACE1* insertion/deletion and *ACE2*rs2285666 polymorphism were determined. Variables frequencies were compared between the groups by logistic regression. We also sequenced the ACE2 coding nucleotides in a group of patients.

**Results:** Severe COVID19 was associated with hypertension male gender (p<0.001), hypertension (p=0.006), hypercholesterolaemia (p=0.046), and the ACE1-DD genotype (p=0.049). In the multiple logistic regression hypertension (p=0.02, OR=2.26, 95%CI=1.12-4.63) and male gender (p=0.002; OR=3.15, 95%CI=1.56-6.66) remained as independent significant predictors of severity. The *ACE2* polymorphism was not associated with the disease outcome. The *ACE2* sequencing showed no coding sequence variants that could explain an increased risk of developing COVID19.

**Conclusions:** Adverse outcome of COVID19 was associated with male gender, hypertension, hypercholesterolemia and the *ACE1* genotype. The *ACE1*-I/D was a significant risk factor for severe COVID19, but the effect was dependent on the hypertensive status.

## Introduction

The SARS-Cov-2 responsible for the COVID19 pandemic is an Angiotensin I converting enzyme 2 (ACE2)-tropicvirus. Like with the SARS-Cov, the “spike” (S) protein of the new beta-coronavirus binds to the nasopharyngeal mucosa and alveolar pneumocytes that express ACE2 at their surface (**1**,**3**). The clinical spectrum of this disease, termed COVID19, ranges from mild to very severe cases (**4**,**5**). **I**t has been hypothesized that viral infection drives an exacerbated inflammatory response, leading to severe lung injury that may require ICU admission, mechanical ventilation and increases the risk of multi-organ failure and death (**6**).

The Renin-Angiotensin-Aldosterone system (RAAS) seems to play an important role in the pathogenesis of COVID19 (**7**). The angiotensin-converting enzyme (ACE1)catalyzes the synthesis of Angiotensin-II (Ang-II) from Ang-I, and ACE2 hydrolyzes Ang-IIinto Ang-1-7. Ang-II binds to the AT1-receptor driving vasoconstriction, fibrosis, inflammation, thrombosis, among other responses; while Ang-1-7 binds to the AT2-receptor with increased vasodilation and reduced fibrosis, inflammation, and thrombosis. The ACE1 and ACE2 are thus seen as opposite players in the balance that determines the risk of developing hypertension and cardiovascular disease. In the lung, ACE2 drives aprotective response by reducing oedema, permeability and pulmonary damage (**8-11**). Of note, hypertension and cardiovascular disease are frequent comorbidities in COVID19, and are strongly associated with the risk of hospitalization and death in individuals exposed to SARS-Cov-2 (**12-15**) (**Suppl. figures**).

Both, acquired and inherited factors associated with differences in the expression and function of the RAAS components could explain the risk of developing COVID19 and its adverse events. For instance, ACE2 expression in the lungs markedly decreases with age and is greater in men than in women (**16**). This could explain the higher risk for adverse outcomes in elderly and male. In general, conditions related with a reduced ACE2 expression would increase the risk for hypertension, cardiac hypertrophy, and heart failure (**17**). In opposition, a high activity of ACE1 would increase the risk of lung and cardiovascular disease by increased activity of the Ang-II/AT1R axis (**18**). Common variants in the two *ACE* genes have been associated with the risk of hypertension, heart disease, renal failure, and pulmonary disease. In fact, the *ACE1* insertion/deletion (I/D) is one of the best characterised human polymorphisms.

Individuals with a D/D genotype showed the highest blood ACE1 levels, and this increased expression would explain the higher risk for cardiovascular and respiratory disease among individuals who are deletion-homozygous. This polymorphism has been related with the outcome in acute respiratory distress syndrome (ARDS) by some authors, and also with the progression of pneumonia in SARS (**19-22**).

The *ACE2* gene is on chromosome X and several single nucleotide polymorphisms (SNPs) have been investigated as risk factors for hypertension and heart failure, including a G to A change at nucleotide +4 of intron 3 (SNP rs2285666) (**23-26**). The fact that *ACE2* is on chromosome X has been seen as a disadvantage for male carriers of alleles linked to a lower ACE2 expression, and could explain the higher prevalence of severe COVID19 among males. SARS-CoVdown-regulate myocardial ACE2 expression and this could explain the myocardial inflammation and damage and adverse cardiac outcomes in patients with SARS (**27**).

Our current knowledge supports a role for the ACE1/ACE2 imbalance in the pathogenesis of COVID19. In this context, variants at these genes associated with differences in gene expression and protein function might explain the individual’s predisposition to manifest the disease symptoms and the risk for hospitalization and adverse events. Moreover, some authors have hypothesised that regional differences in allele frequencies could explain the different rate of the incidence and mortality (**28**,**29**). Our purpose was to determine whether two common functional *ACE1* and *ACE2* variants were associated with susceptibility and outcome in COVID19.

## Patients and methods

### Study cohorts

We collected the anthropometric and clinical data of 204 patients who required hospitalization due to COVID19 (mean age 64.77 years, range 24-95). All the study participants were Caucasian from the region of Asturias (Northern Spain, total population 1 million), and positive for SARS-Cov-2 (PCR test from nasal swabs or tracheobronchial aspirates). Severe cases (n=53) were defined as those in need of critical care support, including high-flow oxygen, positive-pressure ventilation (either invasive or non-invasive) or vasoactive drugs. We also studied 536 healthy population controls matched with the patients for age (n=536; mean age 70.01 years, range 50-81). The presence of comorbidities (hypertension, diabetes, hypercholesterolaemia) was obtained from the participants medical records. The study was approved by the Ethics Committee of Principado de Asturias (Oviedo, Spain). All the patientsor their representatives gave their consent to participate.

### Genotyping

The I/D polymorphism (rs4646994) in intron 16 of the *ACE1* gene was genotyped by polymerase chain reaction (PCR) followed by agarose gel electrophoresis to visualise the two alleles, as reported (**30**,**31**). For the *ACE2*rs2285666 A/G SNP the PCR fragments were digested with the restriction enzyme *AluI* and electrophoresis on agarose gels (**Suppl. figures**).

### *ACE2* sequencing

The *ACE2* coding exons of 60male patients (30 severe and 30 non-severe) were amplified with primers designated from exon flanking introns (**suppl. tables**). These fragments were sequenced with Sanger BigDye chemistry in a capillary ABI3130xl equipment, and the sequences for each patient compared with the ACE2 reference sequence (www.ensembl.org).

### Statistical analysis

All the patients and controls data were collected in an excel file and following the requirements of the Ethical Committee. The statistical analysis was performed with the R-project free software (www.r-project.org). The logistic regression (linear generalized model, LGM) was used to compare mean values and frequencies between the groups.

## Results

Compared to age-matched controls, patients with COVID19 did not differ in the frequency of diabetes, hypertension, and the *ACE*-DD genotype (**table 1**). We found a non-significantly lower frequency of hypercholesterolemia in the patients. We compared these variables in severe COVID19 (patients who required mechanical ventilation and/or ICU supportive care) and mild-disease patients. Male sex, hypertension, hypercholesterolemia, and the *ACE*-DD genotype frequencies were significantly higher in the severe group (**table 1**). The multiple logistic regression-LGM showed that hypertension (p=0.02, OR=2.26, 95%CI=1.12-4.63) and male gender (p=0.002; OR=3.15, 95%CI=1.56-6.66) remained as independent significant predictors of severity.

**Table 1.** Main clinical values and genotype frequencies in total patients and controls.

|  | **Severe**<br>**Disease**<br>**N=67** | **Mild**<br>**Disease**<br>**N=137** | **p** | **All**<br>**COVID**<br>**N=204** | **Controls**<br>**N=536** | **p** |
| --- | --- | --- | --- | --- | --- | --- |
| **Mean age** | 65.76 | 64.56 | 0.53 | 64.96 | 69.88 | AV |
| **Male** | 53 (79%) | 72 (53%) | <0.001 | 125 (61%) | 248 (46%) | AV |
| **Hypertensives** | 41 (61%) | 56 (41%) | 0.006 | 97 (48%) | 228 (43%) | 0.22 |
| **Diabetes** | 15 (22%) | 21 (15%) | 0.21 | 36 (18%) | 123 (23%) | 0.13 |
| **Hyperchols** | 29 (43%) | 40 (29%) | 0.046 | 69 (34%) | 220 (41%) | 0.07 |
| ***ACE I/D*** |  |  |  |  |  |  |
| **DD** | 31 (46%) | 44 (32%) | 0.049* | 75 (37%) | 195 (36%) | 0.92* |
| **ID** | 31 (46%) | 76 (56%) |  | 107 (52%) | 256 (48%) |  |
| **II** | 5 (8%) | 17 (12%) |  | 22 (11%) | 85 (16%) |  |
| ***ACE2#*** |  |  |  |  |  |  |
| **A-carriers** | 52 (78%) | 94 (69%) | 0.18 | 146 (72%) | 374 (75%) | 0.33 |
| **G** | 15(22%) | 43 (31%) |  | 58 (28%) | 124 (25%) |  |

AV: adjust variable, #498 controls genotyped, A-carriers=maleA and female AA+AG.
\* DD vs ID+II

We then compared the values in men and women. In men, hypertension, hypercholesterolemia and the *ACE*-DD were significantly increased in severe patients (**table 2**). The *ACE2* rs2285666 alleles did not differ between the two patients groups and were non-significantly higher to the control frequencies. The same analysis performed among female patients revealed no differences between the severe and mild cases, although we observed a trend toward a higher risk for severity among hypertension and hypercholesterolaemia, and a non-significantly higher frequency of the A-allele in the two patients groups compared to controls (**table 3**). The results in this female cohort would be limited by the reduced size of the severe cohort (n=14).

**Table 2.** Values in the male patients and controls.

| **MALE** |  |  |  |  |  |  |
| --- | --- | --- | --- | --- | --- | --- |
|  | **SEVERE**<br>**N=53** | **MILD**<br>**N=72** | **p** | **COVID**<br>**N=125** | **CONTROL**<br>**N=248** | **p** |
| **Mean age** | 67.19 | 63.89 | 0.13 | 64.77 | 70.01 | AV |
| **Hypertensives** | 34 (64%) | 26 (36%) | 0.002 | 60 (48%) | 97 (39%) | 0.10 |
| **Diabetics** | 12 (23%) | 12 (17%) | 0.42 | 24 (19%) | 47 (19%) | 1 |
| **Hiperlichols** | 25 (47%) | 18 (25%) | 0.01 | 43 (34%) | 82 (33%) | 0.79 |
| **ACE2#** |  |  |  |  |  |  |
| **G** | 43 (81%) | 58 (81%) | 0.92 | 101 (81%) | 200 (85%) | 0.25 |
| **A** | 10 (19%) | 14 (19%) |  | 24 (19%) | 34 (15%) |  |
| **ACE I/D** |  |  |  |  |  |  |
| **DD** | 28 (53%) | 25 (35%) | 0.043* | 53 (42%) | 85 (34%) | 0.13 |
| **ID** | 23 (43%) | 43 (60%) |  | 66 (53%) | 123 (50%) |  |
| **II** | 2 (4%) | 4 (5%) |  | 6 (5%) | 40 (16%) |  |

AV: adjust variable, #234 male controls genotyped.
\* DD vs ID+II

**Table 3.** Values in the female patients and controls.

| **FEMALE** |  |  |  |  |  |  |
| --- | --- | --- | --- | --- | --- | --- |
|  | **SEVERE**<br>**N=14** | **MILD**<br>**N=65** | **p** | **COVID**<br>**N=79** | **CONTROL**<br>**N=288** | **p** |
| **Mean age** | 60.36 | 65.31 | 0.21 | 63.48 | 69.88 | AV |
| **Hypertensives** | 7 (50%) | 30 (46%) | 0.79 | 37 (47%) | 131 (45%) | 0.82 |
| **Diabetics** | 3 (21%) | 9 (14%) | 0.48 | 12 (15%) | 76 (26%) | 0.05 |
| **Hiperlipemics** | 4 (29%) | 22 (34%) | 0.70 | 26 (33%) | 138 (48%) | 0.02 |
| **ACE2#** |  |  |  |  |  |  |
| **GG** | 9 (64%) | 36 (55%) |  | 45 (57%) | 174 (55%) |  |
| **AG** | 3 (21%) | 25 (38%) |  | 28 (35%) | 81 (36%) |  |
| **AA** | 2 (14%) | 4 (7%) |  | 6 (8%) | 9 (9%) |  |
| **A** | 0.25 | 0.25 | 0.87 | 0.25 | 0.19 | 0.07 |
| **ACE I/D** |  |  |  |  |  |  |
| **DD** | 3 (21%) | 19 (29%) | 0.55* | 22 (28%) | 110 (38%) | 0.09 |
| **ID** | 8 (58%) | 33 (47%) |  | 41 (52%) | 133 (46%) |  |
| **II** | 3 (21%) | 13 (20%) |  | 16 (20%) | 45 (16%) |  |

AV: adjust variable, #264female controls genotyped.
\* DD vs ID+II

To better understand the relationship between the two ACE polymorphisms and hypertension, we compared the genotype frequencies between hypertensives and normotensives in male and female patients and controls (**supplementary table**). In males, the ACE-DD genotype had a higher frequency among hypertensives in the three groups, without statistically significant differences (**figure 1**). The *ACE2*-A allele was significantly increased in the hypertensive controls (21% vs. 10%; p=0.02), with no significant differences betweensevere and non-severe COVID19 cases. In the female cohort, the DD frequencies did not differ between hypertensive and normotensive in the three groups. The rs2285666 A allele was significantly more frequent among the hypertensive controls (p=0.04). These results suggested that the *ACE*-DD genotype and *ACE2*-A carriers could be at a higher risk for hypertension in our population, and genotype and allele differences between the severe and non-severe COVID19 cases could be attribute to their association with hypertension.

**Figure 1.**
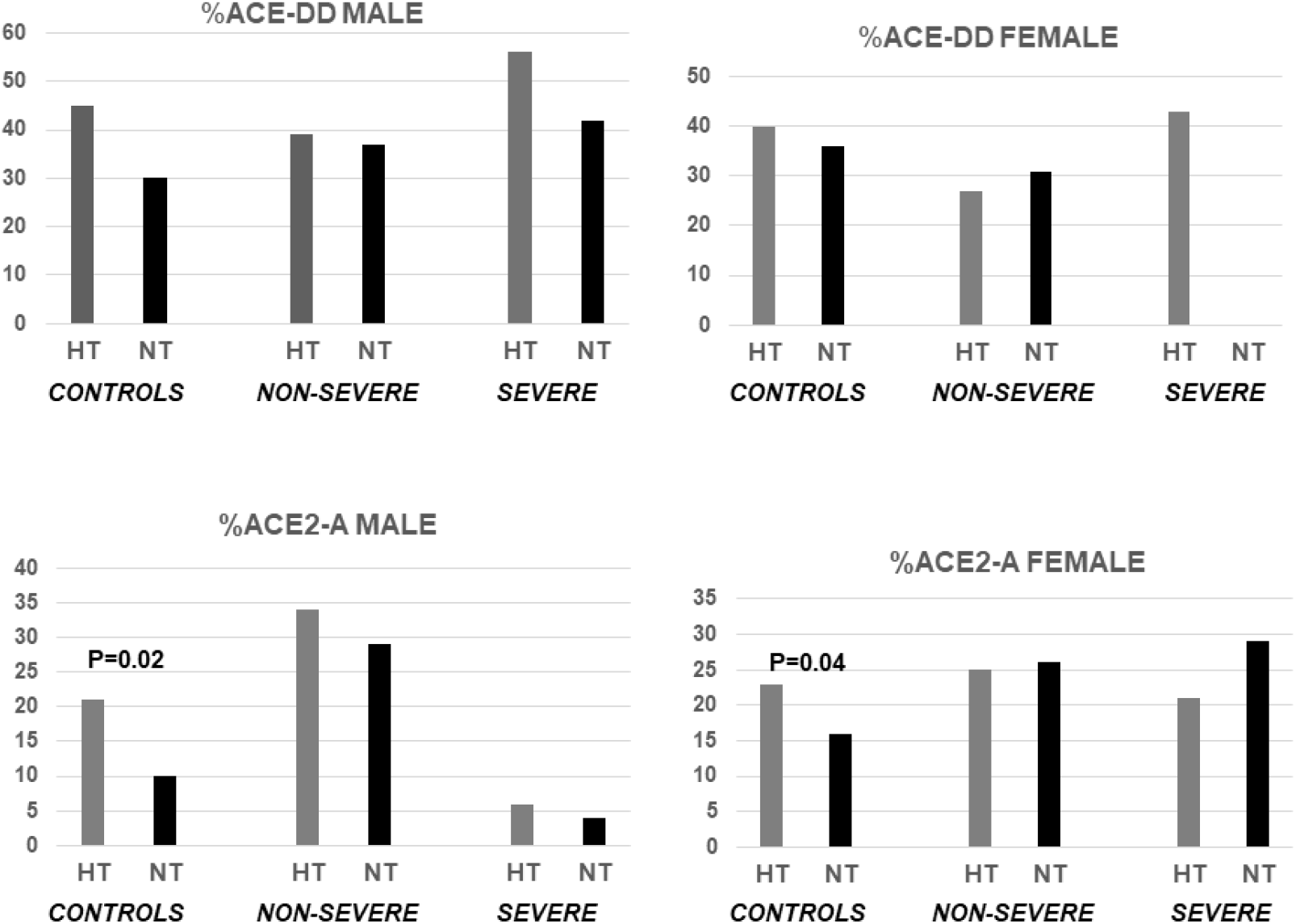
*ACE1* and *ACE2* genotype frequencies according to sex and hypertension.

## Discussion

Since the outbreak of the COVID19 pandemic several authors have speculated about the role of the *ACE1* and *ACE2* gene polymorphisms in disease susceptibility and severity. Because ACE2 is the SARS-Cov-2 receptor, a functional variant that increased gene expression could be associated with a higher number of membrane-bound viral binding sites, increasing the vulnerability of carriers to infection. These risk variants might be particularly adverse in males, who carry only one copy of X-linked *ACE2*gene. The *ACE2* rs2285666 SNP was in the intronic-consensus splicing nucleotides and could thus affect the processing of *ACE2* total RNA to mRNA and, eventually, the amount of the protein. At least one study has reported higher levels of circulating ACE2 levels in men than in women (**32**). One study has investigated the effect of rs2285666 on serum ACE2 levels, and found significantly higher levels in A-carriers compared to G-homozygotes (**33**). A different intronic SNPin strong linkage disequilibrium with rs2285666 (rs879922) has been associated with *ACE2* gene expression (**24**). We thus hypothesised that these functional*ACE2* variants could modify the disease outcome. In our study, the A-allele frequency was non-significantly higher inpatients vs. controls but was associated with hypertension in both, male and female controls. Lower *ACE2* levels should be harmful for patients with lung disease, and the frequency observed in COVID19 patients could thus be the balance between the negative association with viral infection (lower expression in the airway epithelia) and a positive association with respiratory and cardiovascular disease (lower expression in lung and other organs).

The *ACE2* A allele has frequencies of 0.15 and 0.19 in our elderly controls. According to the human gene variation databases, this frequency is lower that the reported among unselected Caucasians (0.20-0.25) including the Spanish population (0.24). This lower frequency could be characteristic of our population, but might also reflect a reduction of the lifespan for rs2285666A carriers.

Functional variants in receptors for other viruses confer resistance to infection. One the best characterised is CCR5, the cellular receptor for HIV. A common variant that determines the absence of the receptor (CCR5-Δ32) confers a complete resistance to HIV-infection among homozygotes, while reduces the disease progression in heterozygotes. Approximately 1% of the Caucasians are *CCR5*-Δ32 homozygotes and they have a non-significant reduction in lifespan (**34**). On the contrary, *ACE2*pathogenic variants are very rare at a population scale, and the complete absence of the receptor would be incompatible with life in humans. Moreover, according to the human genome variation databases, there are no common missense changes in the coding *ACE2* sequence. At a minor allele frequency >0.01% only four missense changes have been reported (all with global frequencies <1%), and rs2285666 was the only variant that could affect splicing (**suppl. tables**). We sequenced the exon and intron-flanking sequence in 60 patients and rs2285666 was the only identified variant. Therefore, it is unlikely that the *ACE2* coding variants have a significant effect on susceptibility to SARS-Cov-2 infection. Of course, this does not exclude that variants in other gene regions are related with gene expression and the amount of protein.

ACE2 expression is regulated by Ang-II through the AT1R-pathway (**35, 36**). ACE1 activity could thus modulate the ACE2 expression and activity through the regulation of Ang-II levels. In this scenario the *ACE2* gene might be down-regulated in *ACE1*-DD homozygotes, who have increased ACE1 activity. There are two mechanisms by which functional *ACE1* variants could modulate the risk of develop and the clinical outcome of SARS-Cov-2 infection. The reduction of *ACE2* expression could protect against viral infection but would also reduce the beneficial effect of *ACE2* in the lung and other organs. At the same time, *ACE1* would enhance the deleterious Ang-II/AT1R response. The *ACE1*-DD has been associated with and increased risk of respiratory distress system by some authors, but not confirmed by others (**20-22**). This polymorphism was investigated in 44 Vietnamese SARS cases and 103 healthy controls who had been exposed to SARS-Covand 50 controls without contact with SARS patients (non-exposed) (**22**). There were no significant differences for DD-frequency among the groups, suggesting that this polymorphism has no effect on the risk for SARS-Cov infection. However, the frequency of the D allele was significantly higher in hypoxemic than in the non-hypoxemic patients, and could thus contribute to the progression of pneumonia in SARS. In our study we did not find differences for DD-frequencies between COVID19 and controls, thus confirming the lack of association with the risk of developing COVID19 symptoms. However, we did not study individuals exposed who remained asymptomatic and we could thus not exclude an effect in the resistance to viral infection. We found a significant higher risk for a severe form of COVID19 in males. This association was not found among females, although the number of severe-disease women was very low (n=14). We confirmed that this genotype was associated with the risk for hypertension in our male controls, with a trend for association with hypertension in the patients. Thus, we concluded that the deleterious effect of the *ACE1*polymorphism on COVID19 outcome was likely due to its association with hypertension.

Male gender, hypertension and hypercholesterolaemia were significantly associated with the risk of developing severe COVID19 in our cohort. The association of adverse outcome with hypertension and sex is well documented. Total Cholesterol and lipoprotein particles might regulate the disease outcome by several ways, including the capacity of the virus to enter the cell membrane and the modulation of the immune response. Hypercholesterolaemia has been associated with COVID19 among Chinese. The study was based on 394 mild, 171 severe, and 32 critical patients, and found significantly lower values of total cholesterol and low-density lipoprotein cholesterol (LDL-Chol) among the critical cases (**37**). This suggested a protective role for high lipid values in the progression from mild to severe COVID19. We did not confirm this effect. On the contrary, the frequency of hypercholesterolaemia was significantly higher in the severe cases, particularly among male.

Our study has several limitations, mainly the reduced sample size of the patients and of female severe cases in particular. This limits the statistical interpretation of the significant and non-significant associations. Also, we did not study subjects exposed to the virus who did not show disease symptoms. These individuals would be resistant to SARS-Cov-2 infection and are crucial for the identification of gene variants associated with disease susceptibility.

In conclusion, our study suggested that the *ACE*-ID polymorphism was associated with the risk of developing severe COVID19 depending on the hypertension status. The *ACE2*rs2285666 variant was associated with hypertension in our elderly population, without significant difference between mild and severe COVID19 patients.

## Data Availability

DATA ARE AVAILABLE UPON REQUEST TO THE CA

## Contributorship

All the authors contributed to this work by recruiting the patients and performing the genetic and statistical analysis. JG, GMA and EC wrote the ms. All the authors approved the submission of this ms.

## Competing interests

None of the authors have competing interests related to this work.

## Acknowledgements

This work was supported by a grant from the Spanish Plan Nacional de I+D+I Ministerio de Economía y Competitividad and the European FEDER, grant ISCIII-Red de Investigación Renal-REDINREN RD16/9/5 (EC).

---

**Supplementary table.**
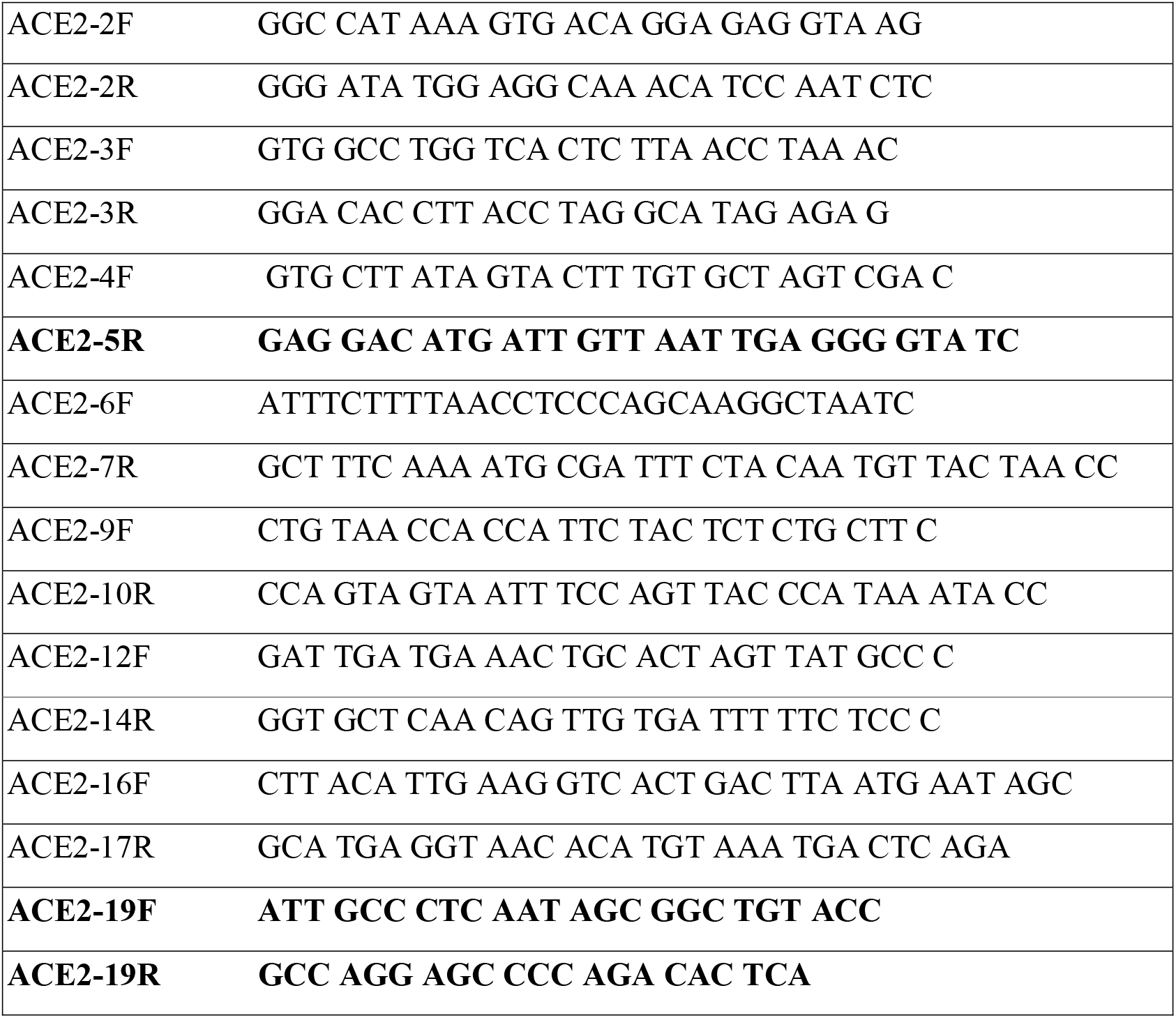
Primers for amplification and sequencing the *ACE2* coding exons.

**Supplementary table.** Distribution of the ACE1 and ACE2 genotypes in patients and controls according to the sex and hypertension.

|  | **MALE**<br>**CONTROLS** |  | **MALE**<br>**MILD** |  | **MALE**<br>**SEVERE** |  |
| --- | --- | --- | --- | --- | --- | --- |
|  | Hypertens<br>N=89 | Normotens<br>N=145 | Hypertens<br>N=26 | Normotens<br>N=46 | Hypertens<br>N=34 | Normotens<br>N=19 |
| **ACE-DD** | 40 (45%) | 44 (30%) | 10 (39%) | 17 (37%) | 19 (56%) | 8 (42%) |
| **ACE-I+** | 49 (55%) | 101 (70%) | 16 (61%) | 29 (67%) | 15 (44%) | 11 (58%) |
| **P**<br>**DD vs I+** | 0.06 |  | 0.88 |  | 0.33 |  |
| **ACE2-A** | 19 (21%) | 15 (10%) | 9 (34%) | 5 (29%) | 6 (17%) | 4 (19%) |
| **ACE2-G** | 70 (79%) | 130 (90%) | 17 (66%) | 12 (71%) | 28 (83%) | 15 (81%) |
| **p A vs G** | 0.02 |  | 0.72 |  | 0.52 |  |

|  | **FEMALE**<br>**CONTROLS** |  | **FEMALE COVID**<br>**MILD** |  | **FEMALE COVID**<br>**SEVERE** |  |
| --- | --- | --- | --- | --- | --- | --- |
|  | Hypertens<br>N=131 | Normotens<br>N=157 | Hypertens<br>N=30 | Normotens<br>N=35 | Hypertens<br>N=3 | Normotens<br>N=19 |
| **ACE-DD** | 53 (40%) | 57 (36%) | 8 (27%) | 11 (31%) | 3 (43%) | 0 |
| **ACE-I** | 78 (60%) | 100 (64%) | 22 (73%) | 24 (69%) | 4 (57%) | 7 (100%) |
| **P**<br>**DD vs I+** | 0.47 |  | 0.67 |  | 0.07 |  |
| **ACE2-AA** | 7 (6%) | 2 (1%) | 1 (3%) | 3 (9%) | 0 | 2 (29%) |
| **ACE2-AG** | 40 (33%) | 41 (29%) | 13 (43%) | 12(34%) | 3 (43%) | 0 |
| **ACE2-GG** | 73 (61%) | 101 (70%) | 16 (54%) | 20 (57%) | 4 (57%) | 5 (71%) |
| **ACE2-A** | 0.23 | 0.16 | 0.25 | 0.26 | 0.21 | 0.29 |
| **p A vs G** | 0.04 |  | 0.92 |  | 0.99 |  |

**Supplementary table.** ACE variants with reported global minor allele frequency (MAF) >0.1%

| SNP id rs | nucleotide | effect | MAF Europeans | MAFSpain |
| --- | --- | --- | --- | --- |
| rs2285666 | c.439+4 G>A | Intron,<br>splicing? | 0.24 | 0.25 |
| rs4646116 | c.77 A>G | p.Lys26Arg | <0.01 | <0.01 |
| rs149039346 | c.2074 T>C | p.Ser692Pro | 0 | 0 |
| rs41303171 | c.2158 A>G | p.Asn720Asp | 0.02 | 0.02 |
| rs147311723 | c.2191 C>T | p.Leu731Phe | 0 | 0 |

**Suppl figure.**
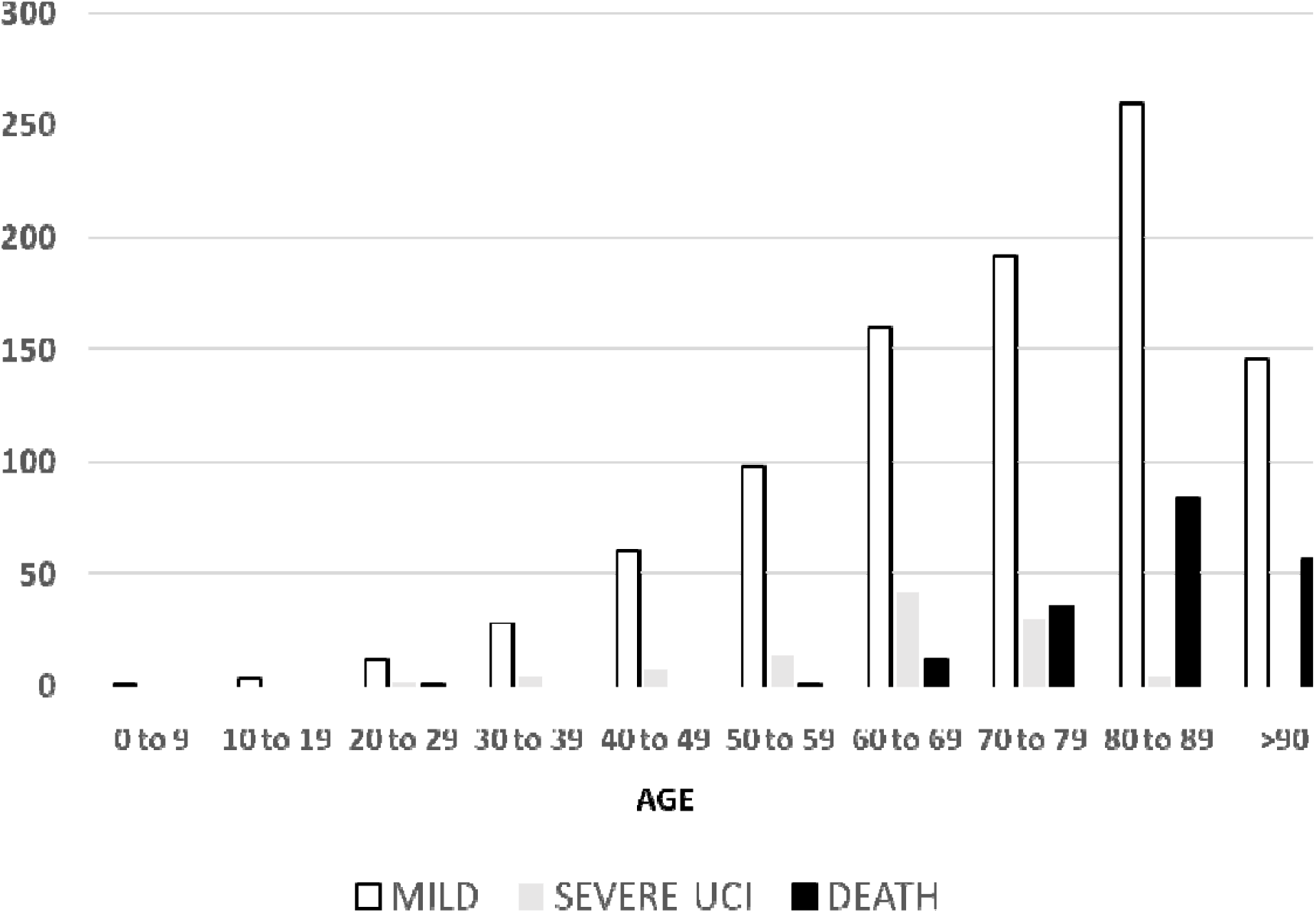
Total number of COVID19 individuals with mild disease, severe-ICU, and deaths in the region of Asturias (population 1 million), according to the age-range. Data released April 14^th^ 2020. Total COVID19 cases, laboratory confirmed: 2,331 (women 1,362). Non-severe hospitalised: 961 (women, 481). Severe-UCI: 103 (women, 31). Death: 191 (women, 103). Source: Asturias Government, https://obsaludasturias.com/obsa/wp-content/uploads/COVID-19_Asturias_Situacion_20200419.pdf

**Suppl figure.**
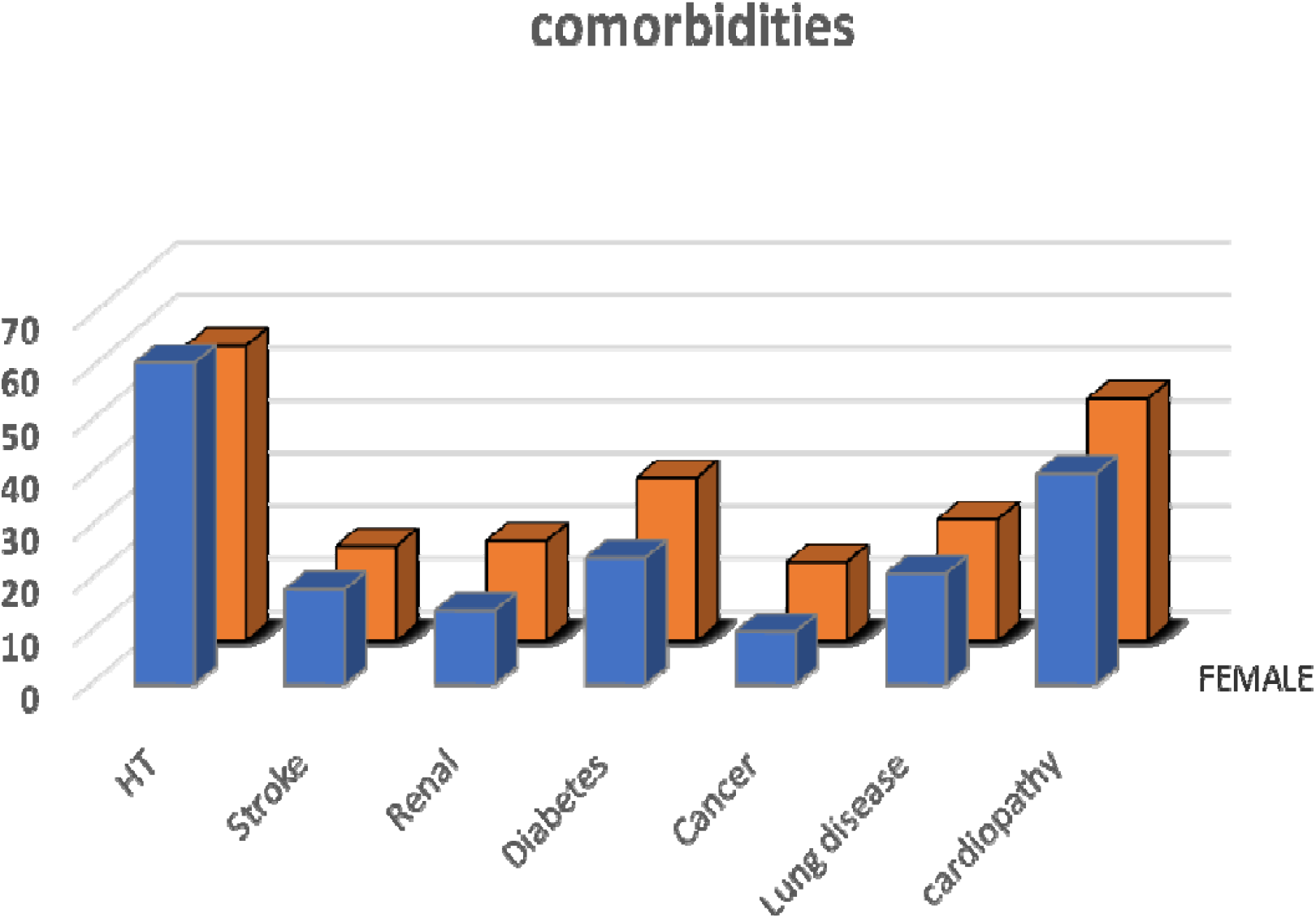
Number of cases with comorbidities among the COVID19 deaths in the region of Asturias, male (blue) and female (orange). Data released April 14^th^ 2020. HT: hypertension, Renal: reduced renal function.

**Suppl. Figure.**
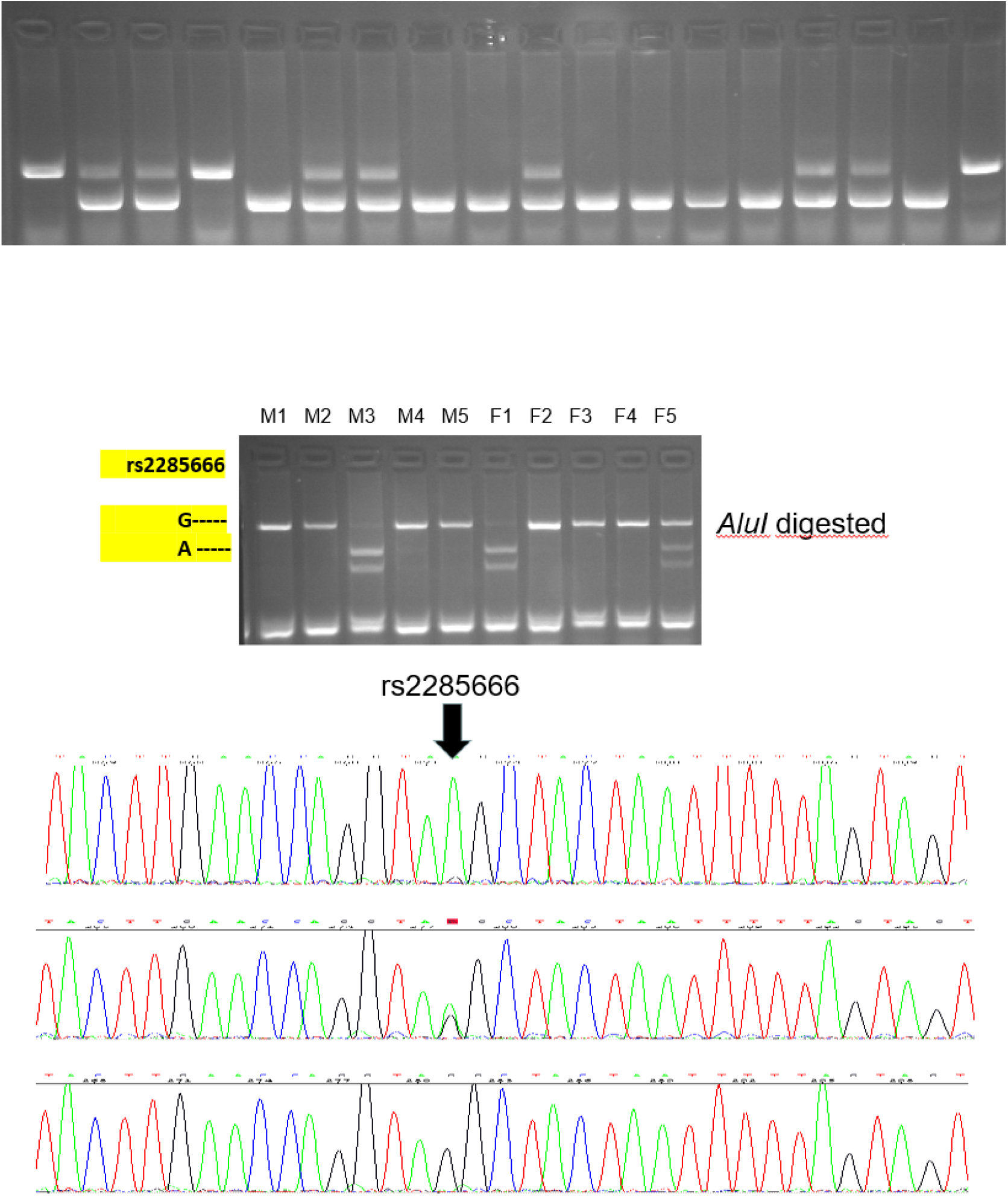
Above: ACE1 AD genotype of 18 individuals. Agarose gel electrophoresis of PCR fragments. Below: Agarose gel electrophoresis of 5 male (M) and 5 female (F) for the ACE2 SNP. Capillary electrophoresis of three *ACE2* genotypes, showing the SNP (arrow).

## Notes

### Competing Interest Statement

The authors have declared no competing interest.

### Funding Statement

This work was supported by a grant from the Spanish Plan Nacional de I+D+I Ministerio de Economia y Competitividad and the European FEDER, grant ISCIII-Red de Investigacion Renal-REDINREN RD16/9/5 (EC).

### Author Declarations

COMITE ETICO DEL HOSPITAL CENTRAL DE ASTURIAS

